# Evaluation of vertical transmission of SARS-CoV-2 in utero: nine pregnant women and their newborns

**DOI:** 10.1101/2020.12.28.20248874

**Authors:** Liang Dong, Shiyao Pei, Qin Ren, Shuxiang Fu, Liang Yu, Hui Chen, Xiang Chen, Mingzhu Yin

**Author notes:** the two authors contribute equally (Liang Dong, Shiyao Pei). **Corresponding Author:** Mingzhu Yin, Hunan Engineering Research Center of Gynecology Disease, Department of Dermatology & Hunan Engineering Research Center of Skin Health and Disease, Xiangya Hospital, Central South University, Changsha 410008, China, Xiang Chen, Department of Dermatology & Hunan Engineering Research Center of Skin Health and Disease, Xiangya Hospital, Central South University, Changsha 410008, China, Hui Chen, Department of Obstetrics and Gynecology, Union Hospital, Tongji Medical College, Huazhong University of Science and Technology, Wuhan 430000, China.

## Abstract

**Background:** Severe acute respiratory syndrome coronavirus 2 (SARS-CoV-2), mainly transmitted by droplets and close contact, has caused a pandemic worldwide as of November 2020. According to the current case reports and cohort studies, the symptoms of pregnant women infected with SARS-CoV-2 were similar to normal adults and may cause a series of adverse consequences of pregnancy (placental abruption, fetal distress, epilepsy during pregnancy, etc.). However, whether SARS-CoV-2 can be transmitted to the fetus through the placental barrier is still a focus of debate.

**Methods:** In this study, in order to find out whether SARS-CoV-2 infect fetus through placental barrier, we performed qualitative detection of virus structural protein (spike protein and nucleoprotein) and targeted receptor protein (ACE2, CD147 and GRP78) expression on the placental tissue of seven pregnant women diagnosed with COVID-19 through immunohistochemistry. Amniotic fluid, neonatal throat, anal swab and breastmilk samples were collected immediately in the operating room for verification after delivery, which were all tested for SARS-CoV-2 by reverse transcriptionpolymerase chain reaction (RT-PCR).

**Results:** The result showed that CD147 was expressed on the basal side of the chorionic trophoblast cell membrane and ACE2 was expressed on the maternal side, while GRP78 was strongly expressed in the cell membrane and cytoplasm. The RT-PCR results of Amniotic fluid, neonatal throat, anal swab and breastmilk samples were all negative.

**Conclusions:** We believed that despite the detection of viral structural proteins in the placenta, SARS-CoV-2 cannot be transmitted to infants due to the presence of the placental barrier.

**Summary:** Our results showed that, excluding environmental pollution after birth and vaginal infection during childbirth, SARS-CoV-2 was less likely to be transmitted vertically in utero.

## Introduction

SARS-CoV-2 is an enveloped single positive stranded RNA virus, which has caused a worldwide pandemic for its high infection rate. It has been proved that SARS-CoV-2 was mainly transmitted by droplets and close contact. Clinically, mild patients have fever, dry cough, and fatigue as the main manifestations, while severe and critically ill patients may have breathing difficulties, acute respiratory distress and other symptoms that are life-threatening [1]. The infection of pregnant women with COVID-19 has been widely concerned since the previous epidemics of emerging viral infections have caused many poor obstetric outcomes including maternal-fetal transmission of the virus, perinatal infections and death, maternal morbidity and mortality. According to the current case reports and cohort studies, the symptoms of pregnant women infected with SARS-CoV-2 were similar to normal adults and may cause a series of adverse consequences of pregnancy (placental abruption, fetal distress, epilepsy during pregnancy, etc.) [1, 2]. However, whether SARS-CoV-2 can be transmitted to the fetus through the placental barrier is still a focus of debate.

Vertical transmission was defined as the transmission of the infectious pathogen from the mother to the fetus during the antepartum and intrapartum periods, or to the neonate during the postpartum period via the placenta in utero, body fluid contact during childbirth, or through direct contact owing to breastfeeding after birth [3]. There were many reports about fetal infection with COVID-19 [1, 2, 4-6], but these experiments did not clarify the relationship between the viral load of placental tissue, amniotic fluid, umbilical cord and neonatal. In addition, there was no distinction between transplacental or transvaginal route and environmental exposure in the experimental design. We retrieved only three case reports and one cohort study, which reported neonatal infection with SARS-CoV-2, that the throat swabs of positive newborns and the placental tissue or amniotic fluid of pregnant women were tested for the virus [7-10]. Two of the 22 newborns delivered by COVID-19 positive pregnant women tested positive for COVID-19 based on the polymerase chain reaction (PCR) results of an NP swab [8]. One case of placental tissue on the maternal and fetal side was positive (RT-PCR), but the fetus had only intrauterine distress and negative for SARS-CoV-2 [9]. Vivanti and his colleagues reported a case of SARS-CoV-2 infection in a pregnant woman who gave birth to a newborn. Amniotic fluid, placenta, neonatal blood and non-bronchoscopic bronchoalveolar lavage fluid were positive for E and S genes of SARS-CoV-2. The neonatal NP and rectal swabs were positive at 1h, 3d and 18d after delivery [7]. According to the published classification for the case definition of SARS-CoV-2 infection in pregnant women, fetuses and neonates, detection of the virus by PCR in umbilical cord blood or neonatal blood collected within first 12 hours of birth or amniotic fluid collected prior to rupture of membrane is considered to be a congenital infection of newborns [11].

The presence of targeted receptors on cell membrane is a prerequisite for virus entry into host cells. Similar to SARS-CoV, SARS-CoV-2 mainly utilizes spike (S) protein to target angiotensin-converting enzyme 2 (ACE2) on the cell membrane, and then entry into the host cell [12, 13]. CD147, also known as Basigin or EMMPRIN, is a transmembrane glycoprotein that belongs to the immunoglobulin superfamily [14]. It is an important marker in tumorigenesis and development, and also plays a role in the infection process of HIV, measles, SARS-CoV and other viruses [15-17]. Recently, studies have shown that CD147 mediates virus entry into cells by interacting with the spike protein of SARS-CoV-2 [18, 19]. The Glucose Regulating Protein 78 (GRP78) or Binding immunoglobulin protein (BiP) is the master chaperone protein of the unfolded protein response (when unfolded or misfolded proteins accumulate) [20-22]. Under normal circumstances, GRP78 is located in the lumen of the Endoplasmic Reticulum (ER). When the cell is stressed, the over-expressed GRP78 escapes the retention of the endoplasmic reticulum and transfers to the cell membrane, which makes it a target for many viruses to enter the cell [23]. Previous studies have shown that GRP78 acts as a coordinating factor to promote the virus’s entry into host cells by increasing the adhesion of Middle East respiratory syndrome coronavirus (MERS-CoV) to target cells [24]. Ibrahim M. Ibrahim and his colleagues used bioinformatics methods to predict the binding ability of GRP78 to the SARS-CoV-2 spike protein [22].

In this study, we discussed the possibility of SARS-CoV-2 crossing the placental barrier and the role of placental barrier in virus transmission. To this end, we collected the placental tissues of 7 pregnant women diagnosed by nucleic acid testing and 2 negative pregnant women. Real-Time Reverse Transcription Polymerase Chain Reaction Assay (RT-PCR) for amniotic fluid, breast milk, neonatal throat and anal swab was examined, and the protein expression of targeted receptors (ACE2, CD147 and GRP78) and viral structural protein (S protein and N protein) in placental tissues were estimated through immunohistochemical experiment to evaluate the possibility of vertical transmission. We hope that these results will provide valuable clinical data for managing pregnant women with SARS-CoV-2.

## Methods

### Case acquisition

The positive placenta tissue came from the pregnant women who were diagnosed with COVID-19 from Tongji and Union Hospital, Huazhong University of Science and Technology, Wuhan. Two cases of negative placental tissues were obtained as negative controls, tested negative by viral nucleic acid at the same time period. Seven female patients were diagnosed as COVID-19 based on the New Coronavirus Pneumonia Prevention and Control Program (6th edition) published by the National Health Commission of China [46]. Treatments were informed to the patient and consent was obtained. Ethical approval for the study was obtained from the Ethics Committee of Wuhan Union and Tongji hospitals of Huazhong University of Science and Technology. Written informed consent was obtained from each enrolled patient.

### Sample collection and SARS-CoV-2 RT-PCR nucleic acid detection

Maternal throat swab samples from the upper respiratory tract were collected at admission in viral-transport medium. Amniotic fluid, placenta, neonatal throat and anal swab samples were collected immediately after delivery in the operating room. The operation was performed in a negative pressure operating room, and doctors and nurses were well protected. All babies were separated from their mothers immediately after delivery and admitted to the neonatal intensive care unit. Nucleic acid RT-PCR detection of newborn swabs was performed at 0h, 24h, and 48h after birth. Breastmilk samples were also collected for SARS-CoV-2 testing to evaluate the evidence of vertical transmission. We believed that the infection came from the contact with mother’s body fluids or the external environment if the newborn’s nucleic acid test was negative at 0h and became positive after 24h and 48h. The presence of SARS-CoV-2 was detected with the Chinese Center for Disease Control and Prevention (CDC) recommended kit (BioGerm, Shanghai, China), following WHO guidelines for RT-PCR [47]. The primers used have been described previously [40].

### Construction and Synthesis of Viral Monoclonal Protein

Antigen preparation: Recombinant protein expression plasmids with P-peptide, 6×His or other labeling were constructed using recombinant DNA technology to purify target proteins by affinity chromatography. (For protein information, see Supplementary Table S1 and Figure S1)

Immunized animals: Each tumor-associated marker antigen immunized 5 BALB/C mice (male and female) aged 8 ∼ 12 weeks. The dosage of recombinant protein antigen for the first immunization was 50ug for each mouse, followed by the addition of Forster’s complete adjuvant, and then the immunization was enhanced for 2-4 times according to the serum antibody titer (the interval was generally 2 weeks). The dosage of recombinant protein antigen for each immunization was 25ug for each mouse, with the addition of Forster’s incomplete adjuvant. Cell fusion was performed 3 ∼ 4 days after the last immunization, when the serum antibody titer of mice reached its peak value.

Cell fusion: In the preparation of mouse B-cell hybridomas using polyethylene glycol (PEG) fusion cells, the B cells from the spleen cells of immunized mice were mixed with mouse myeloma cells, and only the fusion cells formed by B cells and myeloma cells could form hybridoma cells secreting specific antibodies. The hybrid cells of B cells and myeloma cells could be screened by HAT culture.

Cloning and culture of hybridoma cells. Monoclonal hybridoma cell lines were obtained by several rounds of limited dilution. In the 96-well culture plate, the dilution was generally 0.8 cells/well. According to Poisson method, there should be 36% of the Wells as 1 cell/well. The supernatant was detected by enzyme-linked immunosorbent assay (ELISA) and the positive clones were screened.

Antibody identification and Mass production of antibodies. Monoclonal antibodies were identified by enzyme-linked immunosorbent assay (ELISA) and SDS-PAGE (see Supplementary Figure S2). Mice were inoculated intraperitoneally (BALB/C).

### Immunohistochemistry

Formalin fixed and paraffin embedded the placental tissues of seven positive and two negative pregnant women. Five-micron serial sections were taken for immunohistochemistry. Immunohistochemistry was performed using an automated stainer (Bond-III; Leica Microsystems Bannockburn, IL) with ACE2 Monoclonal Antibody (ab15348 [2ug/mL], abcam), CD147 Monoclonal Antibody (ab15348 [2ug/mL], abcam) and GRP78 Monoclonal Antibody (promab). SARS-CoV-2 nucleocapsid protein and spike protein monoclonal antibodies were produced by our cooperation with ProMab Biotechnologies, the antibody specificity has been verified by SDS-PAGE and elisa.

## Result

### Clinical findings

The clinical information of the nine cases was introduced in Table 1. The gestational age at delivery averaged 39 6/7 weeks (range 38–42 weeks, median 40 weeks). XGp1 to XGp7 were positive pregnant women diagnosed by throat swab virus nucleic acid test, No-1 and No-2 were negative pregnant women as controls. Maternal COVID-19 test was performed peripartum on all cases within an average of 9 days before delivery (range from 4 days to 17 days ante partum). XGp3, XGp7 and No-1 were for cesarean section, and the other cases were for vaginal delivery. XGp1, XGp3 and XGp7 had fever during the perinatal period, and XGp7 also had diarrhea. All newborns were singletons and did well in the early perinatal period. Nine newborns had a 1-minute apgar score of 8, and a 5-minute score of 9. The average fetal heart rate before delivery was 136/min (range from 125/min to 154/min). In Real-Time Reverse Transcription Polymerase Chain Reaction Assay for SARS-CoV-2, all pregnant women’s colostrum, amniotic fluid, and neonatal throat swab specimens (0h, 24h and 48h after birth) were negative.

**Table 1.** Cases clinical features

| Case | GA at delivery<br>(weeks) | Days from | Route of<br>delivery | Maternal | Apgar score<br>(1/5 min) | Fetal<br>Heart rate | RT-PCR for SARS-CoV-2 |  |  |  |
| --- | --- | --- | --- | --- | --- | --- | --- | --- | --- | --- |
|  |  | COVID+ test to |  | COVID-19 |  |  | Newborn's |  |  |  |
|  |  | delivery<br>(time range) |  | symptoms<br>peripartum |  |  | Mother's throat<br>swab (n=9) | Mother's milk<br>(n=9) | throat swab<br>(n=9) | Amniotic Fluid<br>(n=9) |
| XGp1 | 39 | 10-15 | VD | Fever | 8/9 | 135 | Positive | Ne | Ne | Ne |
| XGp2 | 42 | 5-10 | VD | No | 8/9 | 135 | Positive | Ne | Ne | Ne |
| XGp3 | 38 | 0-5 | CD | Fever | 8/9 | 130 | Positive | Ne | Ne | Ne |
| XGp4 | 39 | 15-20 | VD | No | 8/9 | 135 | Positive | Ne | Ne | Ne |
| XGp5 | 40 | 5-10 | VD | No | 8/9 | 125 | Positive | Ne | Ne | Ne |
| XGp6 | 40 | 5-10 | VD | No | 8/9 | 154 | Positive | Ne | Ne | Ne |
| XGp7 | 41 | 5-10 | CD | Fever and<br>Diarrhea | 8/9 | 135 | Positive | Ne | Ne | Ne |
| No-1 | 41 |  | VD | No | 8/9 | 137 | Ne | Ne | Ne | Ne |
| No-2 | 39 |  | CD | No | 8/9 | 140 | Ne | Ne | Ne | Ne |

GA, gestational age; CD, cesarean delivery; VD, vaginal delivery; The newborn has a throat swab at 0h, 24h, and 48h after birth for testing. All newborns were negative at three time points.

### Expression of ACE2, CD147 and GRP78 in placental tissue

It could be clearly seen from Fig. 1A that GRP78 has strong membrane and cytoplasmic expression in chorionic syncytiotrophoblast (ST). There was also scattered GRP78 expression in the cell membrane of chorionic cytotrophoblast (CT) (Fig. 1 A). CD147 was expressed in ST cell membrane with polarity, such that expression was strongest at the membrane adjacent to the CT and villous stroma. In most cases, the expression of CD147 was restricted to the basement membrane side, weaker expression in the maternal side and usually showed a circumferential membranous pattern. No CD147 staining was present in villous stroma (Fig. 1 B). ACE2 is highly expressed in ST membrane and also expressed in the extravillous trophoblast (EVT) in the decidua basalis (Fig. 1 C). It was worth noting that the expression of GRP78, CD147 and ACE2 on the ST cell membrane of placental villi of negative pregnant women were relatively weaker than that of positive pregnant women (Fig. 1D-F).

**Fig.1.**
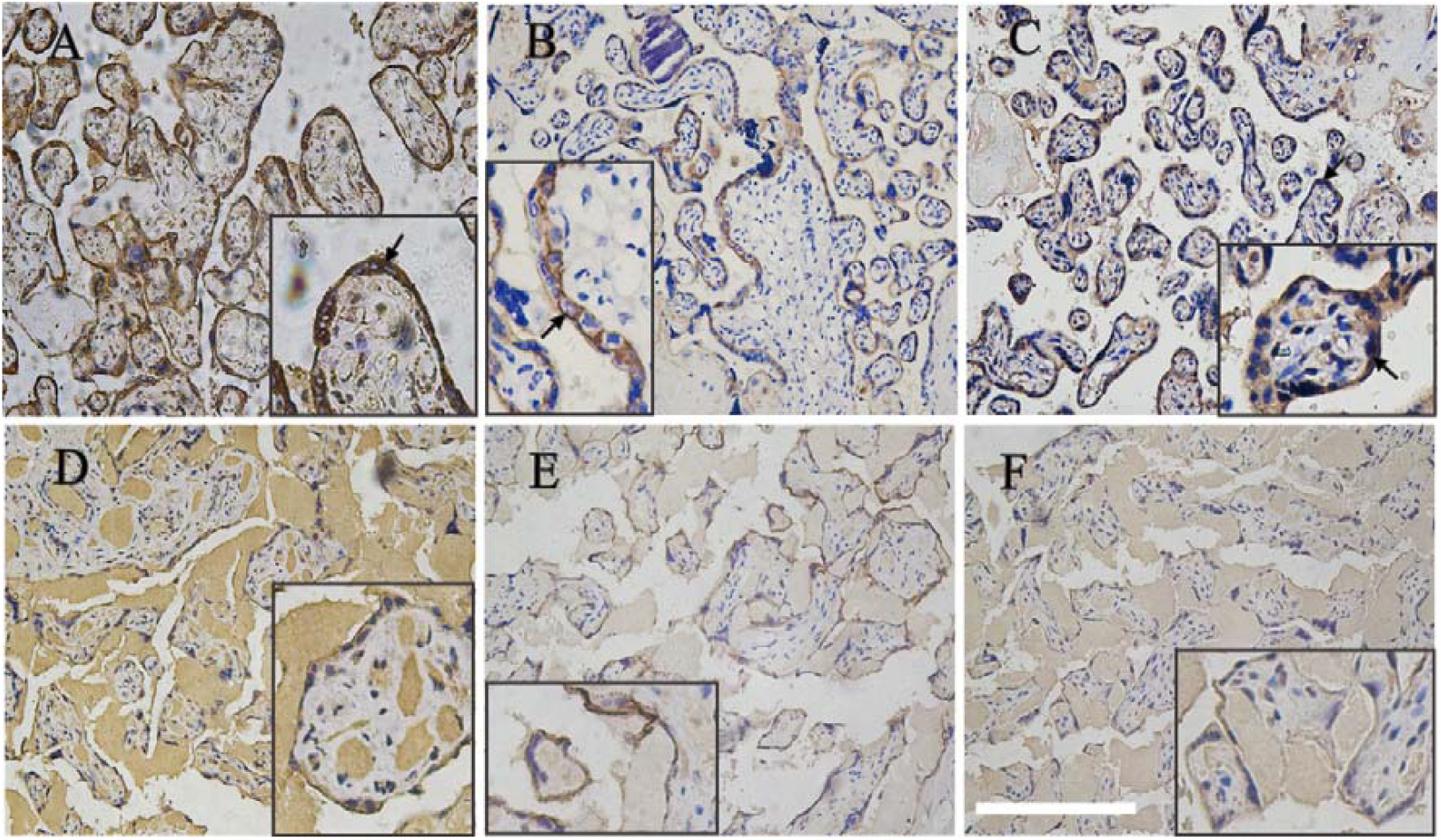
Expression of three host cell receptor proteins of SARS-CoV-2 in placental tissues by Immunohistochemical staining (200×). GRP78 (**A** and **D**), CD147 (**B** and **E**) and ACE2 (**C** and **F**) were all expressed on placental syncytial trophoblasts. **A, B** and **C** came from the placenta tissue of positive pregnant women. **D, E** and **F** were from negative pregnant women. The intense brown cytoplasmic and membranous positivity of syncytiotrophoblast (arrows) (**A**). In our case, ACE2 was expressed on the maternal side membrane of syncytial trophoblasts, while CD147 was expressed on the stromal side (**B**,**C**). The expression of GRP78, CD147 and ACE2 in the placenta tissue of negative pregnant women was relatively weaker than that of positive pregnant women. Bar corresponds to 500um for figs. A-F, 250um for fig. A inset and 200um for figs. B-F inset.

### Viral expression via IHC and Histopathology

Immunohistochemistry of the structural protein of SARS-CoV-2 showed that S protein and nucleoprotein were positive in the cytoplasm of syncytial trophoblasts (Fig. 2). In our research, nucleoprotein was also expressed by rare intervillous macrophages and Hofbauer cells (not shown in the picture). SARS-CoV-2 proteins were not detected in fetal stromal tissue, including villous blood vessels. Pathologic diagnoses were made and lesions graded as per the Amsterdam criteria [25]. Our histopathology result showed that six of the seven positive pregnant women had fibrin deposition between the local villi in the placenta tissue (Fig. 2F). No-1 and No-2 were used as negative controls to verify the specificity of the antibody. The results of two negative controls immunohistochemistry were showed in the supplementary materials.

**Fig.2.**
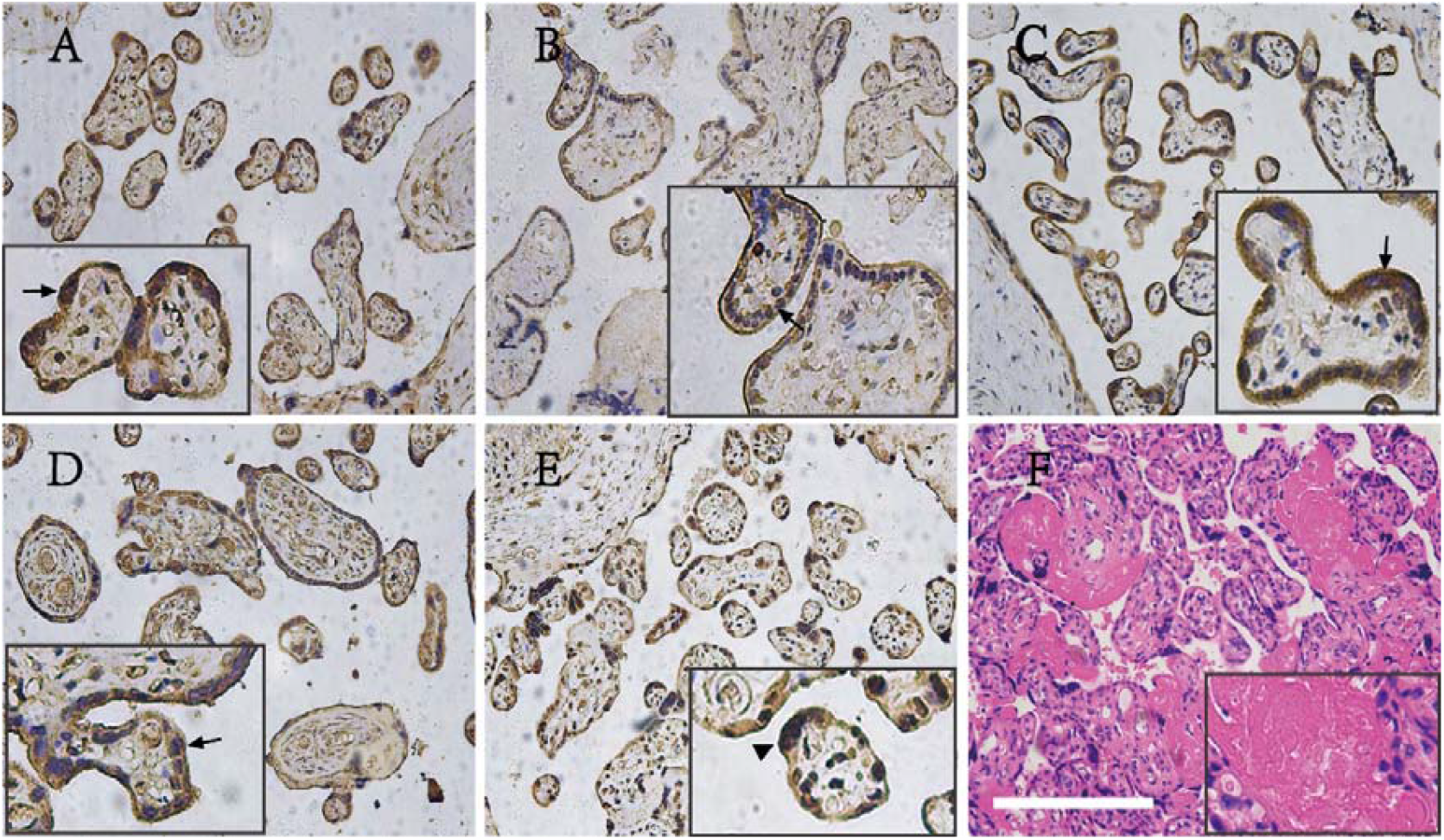
Immunohistochemistry of virus S protein and N protein (200×) and H&E histopathology of COVID-19 exposed placentas (100×). Immunohistochemical stains for His-SARS-CoV-2-NP1 (**A**), His-SARS-CoV-2-NP2 (**B**), His-SARS-CoV-2-NP3 (**C**), SARS-CoV-2-S1(e5)-His (**D**) and His-SARS-CoV-2-S-RBD (**E**). The five viral proteins were all expressed in the cell membrane (arrow). Syncytiotrophoblastic knot shows cytoplasmic staining in **E** (arrowhead). H&E histopathology of COVID-19 exposed placentas (**F**). Bar corresponds to 500um for figs. A-F, 200um for figs. A, C, D and F inset, 250um for fig. B inset and 166um for fig. E inset.

## Discussion

The emergence of SARS-CoV-2 infection has resulted in an epidemic that has rapidly expanded to become one of the most significant public health threats in recent times. Pregnant women and their fetuses represent a potential high-risk population during the COVID-19 outbreak. Given that multiple viruses, such as Varicella-zoster virus, human immunodeficiency virus (HIV), Ebola virus and Zika virus, have been confirmed to be transmitted to the fetus through the placenta and cause a series of adverse pregnancy outcomes (e.g. miscarriage, growth restriction, fetal distress, microcephaly etc.) [26-30]. It is urgently needed to find out whether SARS-CoV-2 infects the fetus through the placenta.

In the study herein, the specimens of the nine pregnant women were collected in the negative pressure operating room immediately after giving birth, and the doctors and pregnant women were well protected. We believed that the specimens collected in this way would minimize the possibility of sample contamination. In addition, throat swabs were collected at 0h, 12h and 48h after delivery. Recently, the SARS-CoV-2 clinical classification of congenital infections in newborns has been published. The possible congenital infections of live births were defined as “detection of the virus by PCR in nasopharyngeal swab at birth (collected after cleaning baby) and placental swab from fetal side of placenta in a neonate born via cesarean section before rupture of membrane or placental tissue” [11]. According to clinical classification, if the fetus has a positive result at 0h and 12h after birth, it can be considered that the infection comes from an intrauterine infection of a COVID-19-positive mother. If it is negative at birth and becomes positive after 48h, it is more likely to be contaminated by the environment. [31]

Currently, the analysis of nasopharyngeal swabs by RT-PCR is the “gold standard” for diagnosing COVID-19 in children and adults, but its efficacy for neonatal diagnosis has not been established. However, many factors affect the sensitivity and specificity of the test and produce laboratory test errors, such as sampling time, sampling method, sample source, storage and transportation, etc. [32]. Studies have shown that the false negative rate of SARS-CoV-2 detected by RT-PCR is related to the exposure time, and RT-PCR analysis may have a higher false negative rate in the early stages of exposure [33]. The Centers for Disease Control and Prevention (CDC) recommends the use of throat swabs as a molecular test because SARS-CoV-2 is mainly transmitted through the respiratory tract and has a high viral load in the nasopharynx [34]. Unlike adults, it is impossible to determine the mechanism of nasopharyngeal virus colonization in infants infected by intrauterine infection. In some cases, specimens collected shortly after birth were negative, and then became positive a few days or even weeks later [8, 35, 36]. On the contrary, there were reports showed that newborns were positive in the early stages [2]. Among our seven newborns with positive mothers, the possibility of false negatives cannot be ruled out, although throat swabs were negative at 0h, 12h, and 48h after birth and the newborns need to be tested and tracked for longer.

Immunohistochemistry technology can locate antigens in placental tissues without destroying the tissue structure and has the advantage of identifying SARS-CoV-2 receptors and nucleic acid expression on specific cell types. The presence of the GRP78, CD147 and ACE2 receptors on the cell membrane is a prerequisite for the placenta to be infected by SARS-CoV-2. In our results, ACE2, CD147 and GRP78 were all expressed in syncytiotrophoblasts. Meanwhile, the immunohistochemical results of the nucleoprotein and S proteins of the syncytial trophoblast virus were positive in the placenta, suggesting that these viruses could infect the placenta and possessed the possibility of crossing the placental epithelial barrier. However, the expression of viral antigens in the villous matrix and interstitial blood vessels, which directly contact with the fetus, was negative. This was consistent with the negative result of our newborn throat swab nucleic acid test. The possible explanation for this result was that the expression of target receptors in the villous matrix was weak, so that the virus could only stayed in the villous syncytiotrophoblast cell layer and cannot enter the villous matrix and interstitial blood vessels to infect the fetus. Roger Pique-Regi and his colleagues performed scRNA-seq studies on placental tissues and found that ACE2 transcription levels were low in placental tissues, which provided a basis for our speculation [37]. Although the immunohistochemical results showed the presence of viral structural proteins in the placental villus syncytiotrophoblasts, the PCR results of neonatal throat and anal swab at 0h, 12h and 48h after birth were all negative, which was indicated that this novel coronavirus was less likely cross the placental barrier to infect the fetus despite it could infect placenta tissues. In addition, the incidence of vertical transmission of SARS-CoV-2 has been considered rare from the limited information. In a meta-analysis, 27 of 936 newborns delivered by COVID-19-positive pregnant women were positive for viral nucleic acid throat swabs, indicating a pooled proportion of 3.2% (95% confidence interval, 2.2-4.3) for vertical transmission [3]. It was noted that the ST membrane of placental tissue of positive pregnant women expressed relatively more receptor proteins than that of negative pregnant women in our study. We speculate that infection with SARS-CoV-2 increases the expression of GRP78, ACE2 and CD147 in ST cell membranes, which makes it easier for the virus to enter syncytiotrophoblasts. However, the underlying mechanism that prevents the virus from crossing the placental barrier needs further studied.

Another possible way for SARS-CoV-2 to be transmitted to infants may be through breast milk. Comparing scRNA-seq datasets extracted from The Cancer Genome Atlas and FANTOM5, it was observed that ACE-2 translation in breast tissue was similar to that of the lung tissue [38]. Despite the presence of ACE2 expression in breast tissue, SARS-CoV-2 was not detected in breast milk in most studies [39-41]. Most guidelines recognize the relative safety of breastfeeding during the COVID-19 period, but more data about breastfeeding safety is still needed [42, 43].

There were many studies reported the abnormal findings in placental pathology among COVID-19-positive mothers. In some cases, HE sections of placental tissue showed low-grade fetal vascular perfusion (FVM). The specific manifestations were vascular occlusion and contiguous, uniformly hyalinized, avascular villi. COVID-19 was often accompanied by a hypercoagulable state of the circulatory system [44], with development of ischemic changes including gangrene of fingers and toes, with evidence of d-dimer elevation, and, in some patients, with disseminated intravascular coagulopathy in one series [45]. However, whether the emergence of FVM is related to the hypercoagulable state associated with COVID-19 needs further research. In our study, we found no specific pathological changes related to COVID-19.

Our advantage lies in performing a comprehensive PCR test and immunohistochemical analysis on the collected specimens. Neonatal swabs have been analysed with paired maternal throat swab, breast milk and amniotic fluid to exclude possible environmental contamination after delivery. However, there were still some limitations in our study, on the one hand, there were only seven cases of positive pregnant women, which might be biased when analyzing the results, on the other hand, we lack cord blood and newborn blood samples. According to the classification of clinical cases, if RT-PCR results are positive in umbilical cord and newborn blood samples, it is even more reasonable to believe that the infection originates from the transmission of the positive pregnant woman to the fetus through the placenta [11].

## Data Availability

All data in this study are true and reliable.

## Acknowledgments

We thank all the hospital staff members for their efforts in collecting the information that used in this study; thank the patients who participated in this study, their families, and the medical, nursing, and research staff at the study centers.

## Authors and contributions

Liang Dong, Shiyao Pei collected the data. Liang Dong and Mingzhu Yin drafted the manuscript. Hui Chen, Xiang Chen contributed to the interpretation of the results and critical revision of the manuscript for important intellectual content and approved the final version of the manuscript. Shuxiang Fu, Liang Yu were involved in data cleaning, and verification. All authors have read and approved the final manuscript.

