## Supplemental table 1 and Supplemental figure 1-3 for "Evaluation of vertical transmission of SARS-CoV-2 in utero: nine pregnant women and their newborns"

**Supplemental Appendix list**

Table S1

Figure S1-3

**Table S1** The information of synthetic virus structural protein

| Protein Name | Expression System | Full Length or Truncated | Protein Description | Purity | Concentration  (mg/ml) |
| --- | --- | --- | --- | --- | --- |
| COVID19-N | E.coli | Full length:1-420aa | COVID-N with His-tagged,  dialyzed against 1×PBS (pH 7.2), soluble  . | 80% | 1 |
| SARS-CoV-2-NP-1 | E.coli | Truncated:1-180aa | COVID19-NP1 with His-tagged,  dialyzed against 1×PBS (pH 7.2), soluble. | 80% | 0.8 |
| SARS-CoV-2-NP-2 | E.coli | Truncated:120-300aa | SARS-CoV-2-NP2 with His-tagged,  dialyzed against 1×PBS (pH 7.2), soluble. | 95% | 25 |
| SARS-CoV-2-NP-3 | E.coli | Truncated: 240-419aa | COVID19-NP3 with His-tagged,  dialyzed against 1×PBS (pH 7.2), soluble. | 85% | 0.5 |
| SARS-CoV-2-S1(e5) | E.coli | Truncated: 523-661aa | SARS-CoV-2-S1(e5) with His-tagged,  dialyzed against 1×PBS (pH 7.2), soluble. | 95% | 0.3 |
| SARS-CoV-2-S-RBD(TV) | E.coli | Tuncated:330-520aa | SARS-CoV-2-S-RBD(TV) with His-tagged,  dialyzed against 1×PBS (pH 7.2), soluble. | 95% | 0.2 |

**Figure** **S1** The recombinant protein sequence

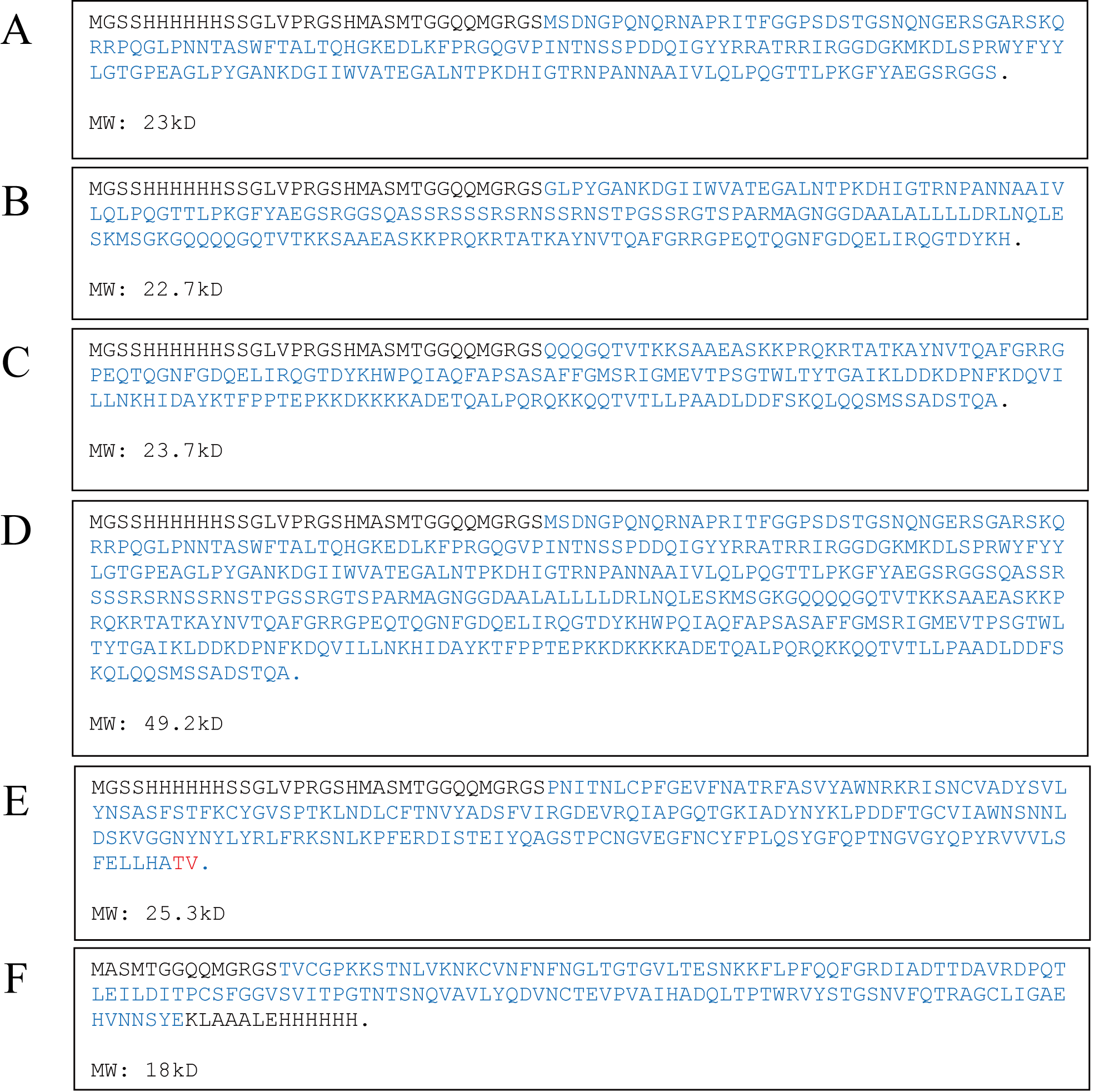

His-SARS-CoV-2-NP1 (**A**), His-SARS-CoV-2-NP2 (**B**), His- SARS-CoV-2-NP3 (**C**), His- SARS-CoV-2-N (**D**), His-SARS-CoV-2-S-RBD(TV) (**E**), SARS-CoV-2-S1(e5)-His (**F**) The protein amino acid sequence is marked in blue font.

**Figure** **S2** The quality of the synthesized viral structural protein was detected by SDS-PAGE and Elisa.

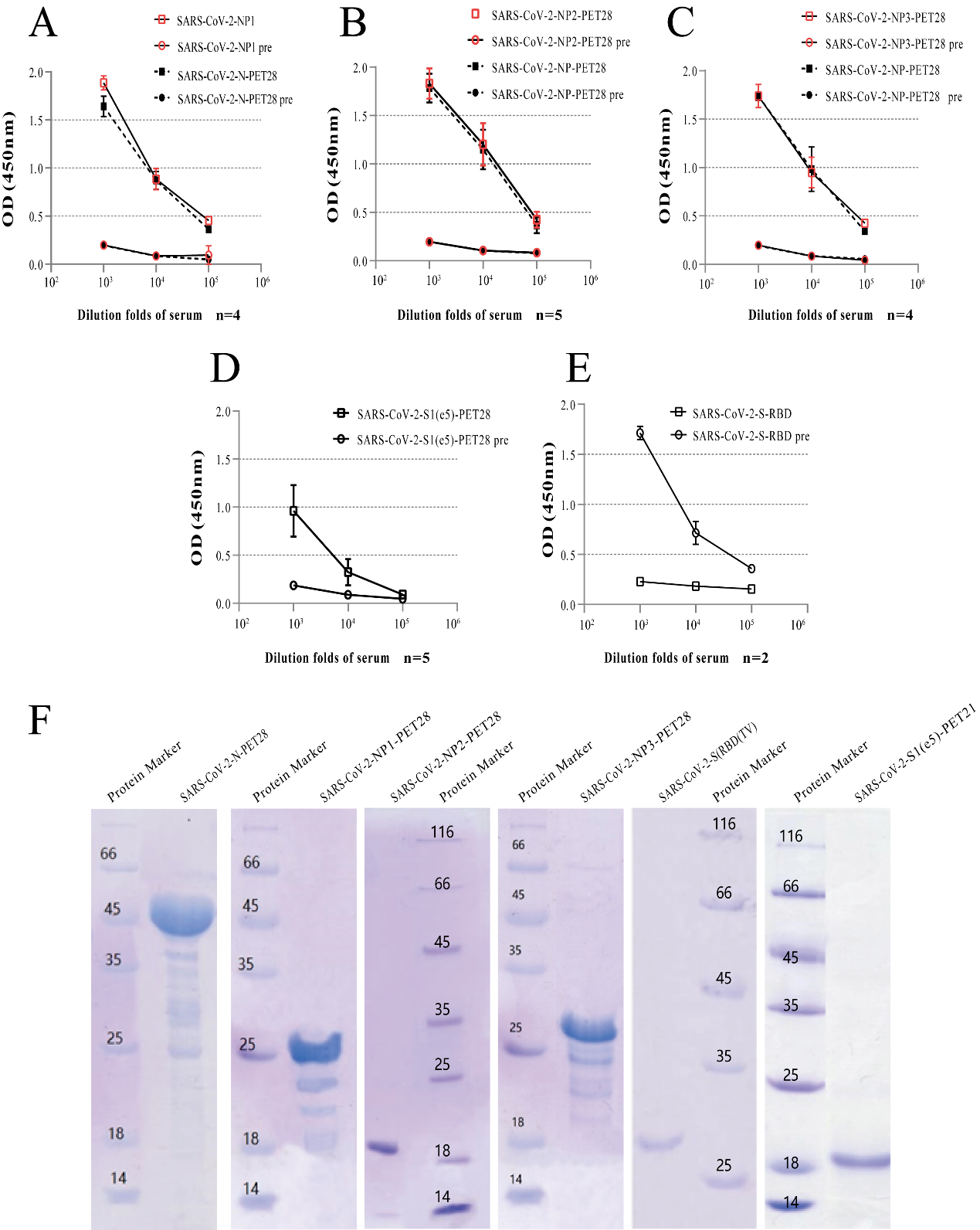

The viral structural protein preparations were used as coating antigen for an elisa. The rabbit or mouse antisera were diluted serially from 1:10^3^ to 1:10^5^. Experiment were repeated several times, and the results were expressed as the mean value (**A**, **B**, **C**, **D** and **F**). The expression of synthetic protein was detected by SDS-PAGE analysis (**F**). n, number; pre, serum harvested from each rabbit or mouse prior to immunization.

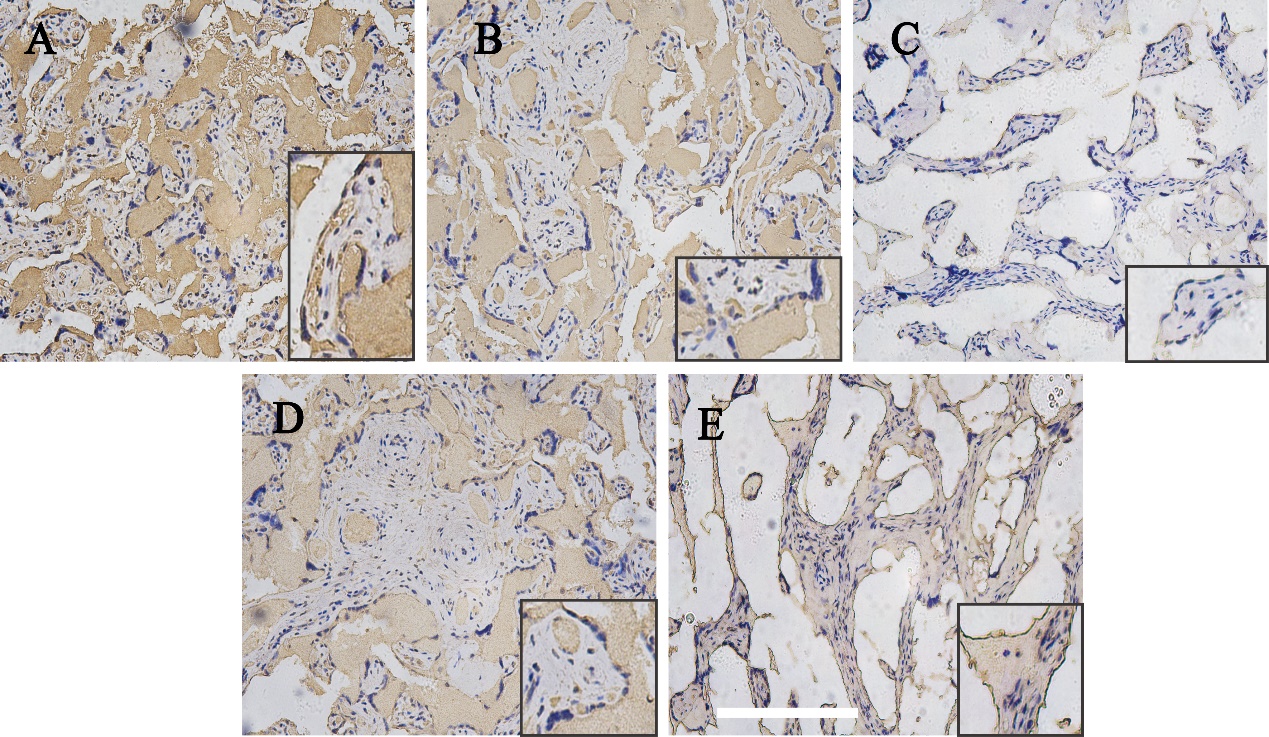

**Figure** **S3** Immunohistochemical expression of virus nucleoprotein and S protein in placenta tissue of two negative patients.

Immunohistochemical stains for His-SARS-CoV-2-NP1 (**A**), His-SARS-CoV-2-NP2 (**B**), His-SARS-CoV-2-NP3 (**C**), SARS-CoV-2-S1(e5)-His (**D**) and His-SARS-CoV-2-S-RBD (**E**). Bar corresponds to 500um for figs. A-E, 250um for figs. A-E inset.
